# Building confidence in crises – the roles of Sierra Leonean religious leaders’ during the 2014-2016 Ebola outbreak

**DOI:** 10.1101/2023.11.14.23298507

**Authors:** Padraig Lyons, Maike Winters, Mohamed F. Jalloh, Helena Nordenstedt, Helle Mölsted Alvesson

## Abstract

**Background:** Religious leaders have been involved in risk communication campaigns for many years across West Africa such as their involvement in the HIV/AIDS pandemic response. Little is known about religious leaders’ perceptions of their role in communicating Ebola risks during the 2014-2016 outbreak in Sierra Leone and the strategies they adopted to communicate these risks.

**Methods:** In this qualitative study 10 semi-structured interviews were conducted with religious leaders in Freetown, Sierra Leone, to better understand their perceptions of their roles communicating risk during the outbreak. Five Christian and five Islamic leaders were recruited from multiple national religious organisations including male and female leaders. The data was analysed using thematic analysis.

**Results:** Three themes were developed from the data which illustrate the different strategies religious leaders used when establishing public confidence in their role communicating risk and the messages they communicated during the outbreak. The religious leaders describe how they established themselves as non-political actors in the outbreak response, they provided support to their communities and used collaboration as a means to effectively communicate risk. The religious leaders adapted pre-existing roles and established new ones in order to build confidence among their communities. They flexibly and pragmatically utilised scriptural texts within these strategies to communicate risk and to establish confidence in their messages.

**Conclusions:** Religious leaders were pragmatic in their approach to risk communication, leveraging political distrust and collaborating with other actors to strengthen their position. Interreligious unity as well as scripturally supported messaging helped establish confidence in the public health emergency response.

**Author Summary:** Religious leaders are both positively and negatively associated with the promotion of protective behaviours during outbreaks. During the 2014-2016 Ebola outbreak, thousands of religious leaders were engaged to communicate Ebola risks and protective Ebola messages across Sierra Leone. Working with public health professionals they developed Ebola specific messages by selecting specific passages from scripture to support medical messages. These messages were positively associated with safe burial behaviours during the outbreak in Sierra Leone.

In this study, semi structured interviews were conducted with religious leaders involved in risk communication campaigns during the outbreak. Our results highlight that religious leaders were pragmatic in their approach to risk communication, leveraging political distrust and collaborating with other actors to strengthen their position. Interreligious unity as well as scripturally supported messaging helped establish confidence in the public health emergency response. In their role communicating risk religious leaders prioritised building confidence by advocating for community supported Ebola-measures and by establishing themselves as non-political actors in the outbreak response.

Public health messages developed by religious leaders in line with scriptural texts can be used as part of risk communication campaigns to improve their public acceptability.

## Introduction

Across the world, religious leaders are trusted figures in their communities and as such, can be positive agents of health behaviour change with involvement in public health interventions (1–4). For instance, hearing messages from religious leaders (pastors or other ministerial staff) that were supportive of same-sex relationships was associated with an increased willingness to use HIV prevention strategies (e.g. pre-exposure prophylaxis) among Black Americans in the United States (5). In Ghana, an intervention in which national and local religious leaders spread HIV public health messages on mass media was associated with reduced HIV-related stigma (6).

Religious leaders have also been associated with hindering public health efforts (7). In the early 2000s in Northern Nigeria, religious leaders propagated misinformation that the oral polio vaccine contained the HIV-virus and antifertility agents. This misinformation contributed to the suspension of the polio vaccine campaigns in the region (8). During the HIV/AIDS epidemic, some religious groups in Nigeria and Burkina Faso opted to promote abstinence over condom use (9, 10), while others actively encouraged condom use to reduce the spread of infection (11). The mixed messages also reflect the influence religious leaders had; they have been both positively and negatively linked to the HIV/AIDS epidemic (6, 9, 10, 12, 13). Religious leaders also have the power to influence vaccine uptake. During a measles outbreak in a Somali community in Minnesota, the engagement of religious leaders helped to increase vaccination rates (14).

The Ebola epidemic that affected West Africa between 2014-2016 posed a unique religious challenge: religious and cultural practices (such as washing a dead body before burial) were strongly associated with Ebola transmission (15, 16). Religious leaders had traditionally promoted various forms of physical contact with corpses including washing of the deceased as part of long-standing funeral and burial rituals (Hewlett 2003).

In order to influence health-behaviours during epidemics and pandemics, risk communication campaigns must appreciate the complex interactions that exist between individuals, their peer-groups and wider communities within a society as described by the socio-ecological model (17, 18). Therefore, a key intervention to improve the uptake of safe burials was to engage religious leaders to promote safe Ebola burials. In collaboration with public health professionals, religious leaders developed scripturally-based messages that were delivered nationwide in places of worship and local community events as well as through radio and television programmes (19, 20). Public health messages promoted by religious leaders were positively associated with willingness to engage in safe burial behaviours during the Ebola outbreak in Sierra Leone (21).

In times of crisis, people will turn to those they trust as a source of information (22). Religious leaders are trusted figures in Sierra Leonean society (23). They were recognized as influential spokespersons for protective health behaviours as part of risk communication campaigns during the Ebola outbreak (24). The COVID-19 pandemic further demonstrated the important role of trusted actors in the public health response, including religious leaders (25). The rampant misinformation surrounding the COVID-19 vaccines made the uptake of protective behaviours challenging in some areas (26, 27). In Romania for example, the strong standing of religion combined with negative attitudes towards vaccines held by some influential religious leaders is thought to have contributed to low vaccination coverage (3). In Israel, on the other hand (a population with very high vaccination coverage) religious leaders were key in community engagement around COVID-19 vaccination (28). Religious leaders were also involved in the COVID-19 response in Sierra Leone including promoting the uptake of vaccinations (29).

Our aim with this study was to understand and contextualise the strategies religious leaders adopted to build confidence and communicate risk during the Ebola outbreak in Sierra Leone.

## Methods

### Setting

Religion plays an important role in the lives of Sierra Leoneans. The majority of Sierra Leoneans affiliate with either Islam (77%) or Christianity (22%) (30). Countrywide, religious leaders are involved in various public health campaigns through faith-based organisations such as the Islamic Action Group (ISLAG) and the Christian Action Group (CHRISTAG). Both these organisations were formed in 1987 with support from the Ministry of Health and UNICEF to undertake social mobilisation activities to promote childhood immunisation. The Action Groups have since been involved in education, water and sanitation, family planning and HIV/AIDS prevention. During the civil war in Sierra Leone (1991–2002) ISLAG and CHRISTAG were brought together to form the Inter-Religious Council of Sierra Leone (IRCSL), an umbrella body responsible for coordination and advocacy on matters involving religious leaders (1). The IRCSL helped coordinate and support dialogue between both parties involved in the conflict and helped facilitate peace negotiations (31, 32). Inter-religious collaboration in Sierra Leone has been described as “a framework of everyday life in Sierra Leone”, capturing the fundamental nature of the collaboration across the country (31, 33). The IRCSL remains actively involved in a wide range of religious, health and development projects, collaborating with government, other faith-based organisations and civil society.

The Religious Leaders’ Taskforce on Ebola was established in July 2014, two months after the first Ebola case was confirmed in Sierra Leone, to enhance the role of faith-based organisations in the response (20). Religious leaders were identified as key figures in community engagement and risk communication efforts. In collaboration with public health professionals, religious leaders developed faith-based messages that were delivered nationwide in places of worship and local community events as well as through radio and television programmes (19, 20).

Religious leaders built trust by engaging the public in dialogue around accurate measures of Ebola prevention. They worked to address people’s concerns and worked to dispel Ebola misinformation in collaboration with public health professionals (20). Messages covered key aspects of Ebola risk communication, drawing upon scriptural texts to promote safe Ebola practices including avoidance of deceased family members and promoting the update of Ebola burial teams (21).

This study was carried out in Freetown, the capital of Sierra Leone where around 1 million of the country’s 8 million people live. The country ranks among the poorest countries on Earth, with one of the highest maternal mortality rates and had a population with a life expectancy of 60 years in 2021 (34).Freetown was one of the worst affected areas during the outbreak with high numbers of positive cases and deaths (35). Participants in this study lived and worked in Freetown at the time of interview. This study was conducted in collaboration with FOCUS1000 (a local NGO) who were one of the organisations involved in the coordination of community engagement and risk communication programmes during the outbreak.

### Study design & participant identification

This study is based on semi-structured interviews with religious leaders that were involved in risk communication campaigns across Sierra Leone during the Ebola outbreak 2014-2016. Ten interviews were conducted with religious leaders in Freetown between March 5^th^ to March 20^th^, 2019. All ten of those invited to interview, attended. Religious leaders were recruited using a combination of purposive and convenience sampling, using the predefined criteria that all participants must have been involved in risk communication activities during the outbreak. FOCUS1000 assisted in recruiting the initial Christian and Islamic interviewees through their local contact networks. Recruitment was conducted with the help of a local Imam who was also interviewed in the study. The sample was predominantly male (n=7) with five Christian and five Islamic leaders included. Five of the leaders were pastors and four were Imams, and the final interviewee was one of the leaders of the National Islamic Council. All the religious leaders held multiple positions across religious organisations in the country, including leadership positions with ISLAG, CHRISTAG and IRCSL. Basic characteristics of the participants in the study are described in Table 1 with their specific roles within faith-based organisations omitted to protect the identity of participants.

**Table 1.** Participant information.

| Interview ID | Gender | Interview length<br>(Mins) | Religious<br>background | Interview location |
| --- | --- | --- | --- | --- |
| Interview 1 | Male | 57 | Islam | FOCUS1000 |
| Interview 2 | Male | 69 | Islam | FOCUS1000 |
| Interview 3 | Male | 43 | Islam | Mosque |
| Interview 4 | Male | 83 | Christian | FOCUS1000 |
| Interview 5 | Female | 25 | Islam | FOCUS1000 |
| Interview 6 | Male | 70 | Christian | FOCUS1000 |
| Interview 7 | Male | 48 | Christian | FOCUS1000 |
| Interview 8 | Female | 74 | Christian | Pastor's home |
| Interview 9 | Male | 57 | Christian | FOCUS1000 |
| Interview 10 | Female | 33 | Islam | FOCUS1000 |

Initial analysis was driven by the WHO guidelines for Risk Communication in Emergencies which provided a framework for analysing risk communication in emergency settings (WHO 2018). The interview guide was largely informed by this framework and a literature search on risk communication strategies during emergencies. Questions were designed in a chronological fashion asking questions related to experiences before, during and after the outbreak. The interview started with open ended questions regarding the participant’s religious background and previous risk communication experience, continuing to questions regarding Ebola knowledge and then more targeted questions regarding experiences conducting risk communication campaigns during Ebola including barriers and facilitators to this process. Finally, open-ended questions were posed regarding reflections on the experience and thoughts going forward. Two pilot interviews were conducted leading to a few minor adjustments to question-wording and ordering before finalising the guide for the remaining interviews.

### Data collection and analysis

All the interviews were conducted in person, in English. The native language of the interviewees was Krio, but all interviewees were fluent and comfortable giving the interview in English. Eight of the ten interviews were conducted in the headquarters of FOCUS1000 (a local NGO) one took place in a local religious school and one in a pastors’ home. PL was in Freetown for one month in 2019 to carry out the interviews with religious leaders as part of a master’s thesis in Global Health. Noone else was present during the interviews which took place at a time chosen by the participants and were conducted both during and after office hours. Interviews lasted between 26 and 83 minutes (Table 1). PL established a relationship with the interviewees via FOCUS1000 and all interviewees were made aware that the research was a part of a masters’ dissertation. Repeat interviews were not conducted and interviews were not returned to participants for comment due to the length of time that elapsed between analysis and conducting of interviews. Brief field notes were made after each interview to summarise any thoughts the lead author relating to questions and responses.

Interviews were transcribed verbatim and analysed in Dedoose, version 8.3.4. Thematic analysis was used to analyse the data, with initial open coding of three of the transcripts using both in vivo codes and codes derived by the lead author (PL). This coding framework was then applied across the remaining 7 interviews. When all transcripts had been coded, several emerging codes were retrospectively applied across the interviews and those first transcripts were revised again to check for consistency in the coding process. The codes were grouped in categories leading to the formation of sub-themes and the final themes. During the course of the analysis, authors discussed and provided feedback on the codes and the emerging themes. In comparing and contrasting the religious leaders’ responses to one another, the analysis became more latent allowing for a deeper understanding of their opinions on their work during the outbreak.

### Ethical considerations

The Sierra Leone Research and Scientific Review Committee granted ethical approval for this study in March 2019. All interviewees provided informed consent to participate by signing an informed consent form.

## Results

Three primary themes were identified from the data representing the strategies religious leaders used to enhance public confidence in their role as risk communicators and the messages they communicated: providing community support, collaboration, and establishing themselves as non-political figures. The religious leaders discuss these three themes as strategies they flexibly used to build confidence in their role communicating risk during the outbreak.

The religious leaders adapted pre-existing roles and established new ones in order to build this confidence during the outbreak. Scripture is ingrained in most aspects of religious leaders’ communication with the public and they flexibly made use of religious texts when establishing confidence in their Ebola messages across these themes. Themes are categorised and summarised in table 2. They are further discussed and illustrated with quotes below.

**Table 2.**
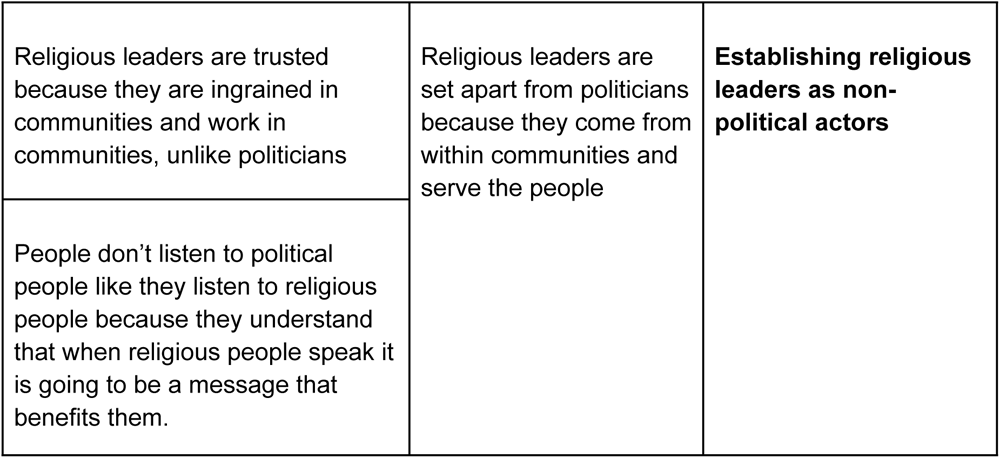
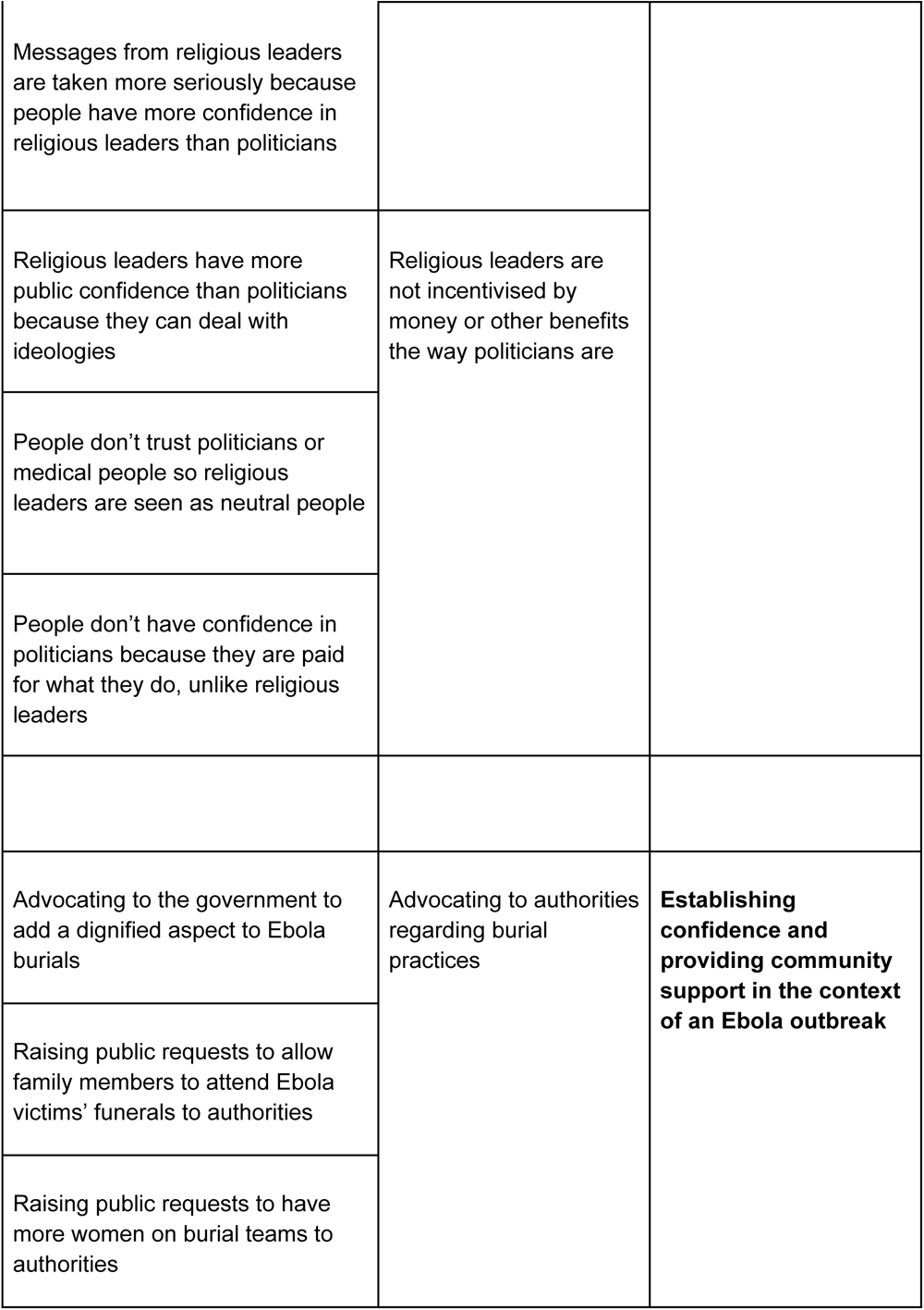
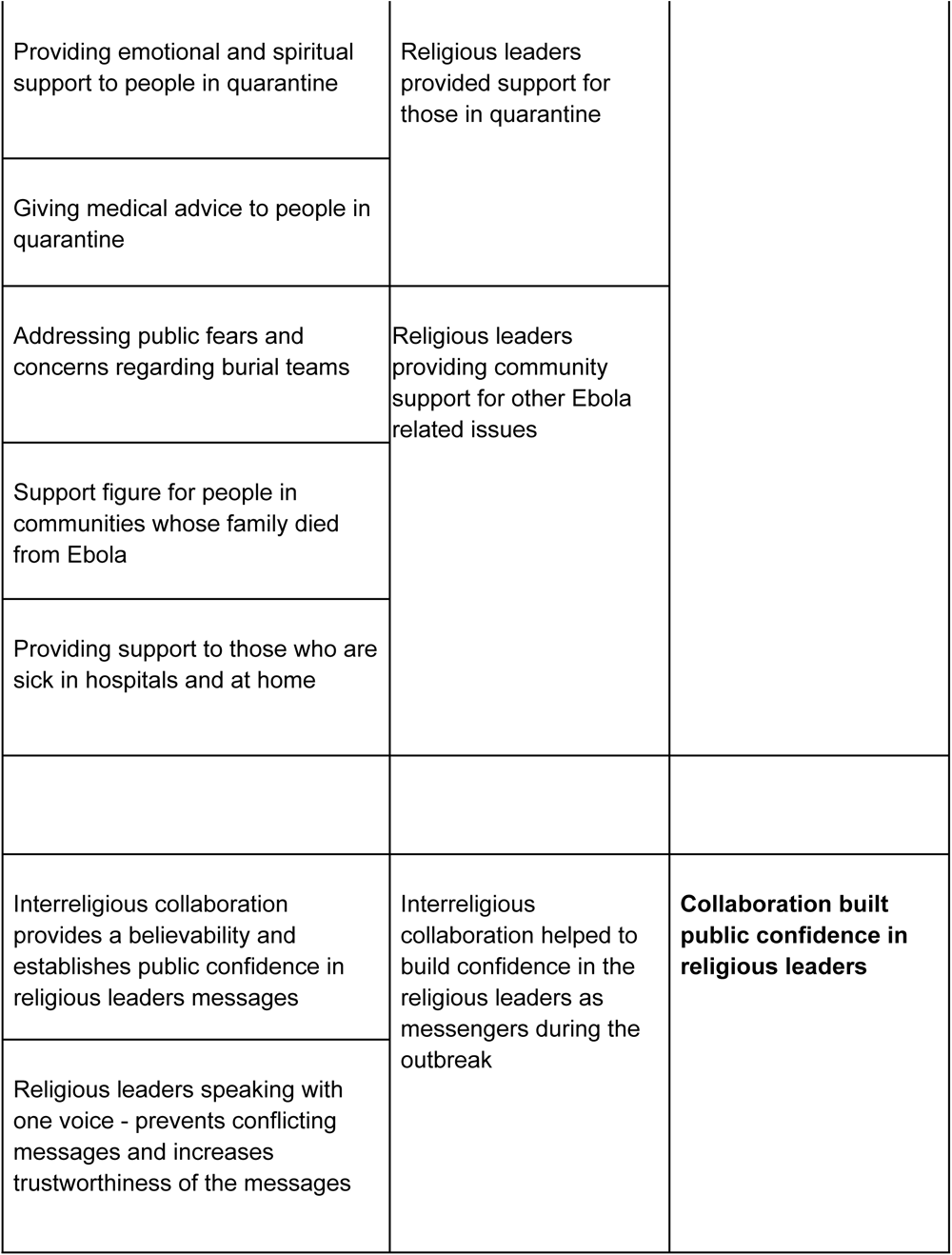
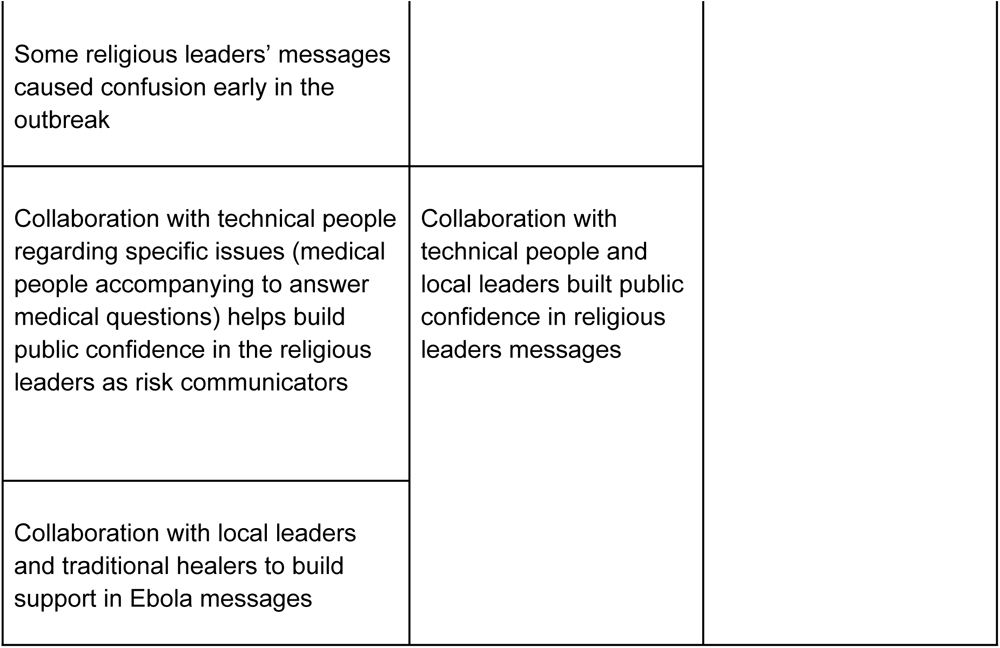
Categories, Sub-themes and Themes.

|  |  |  |
| --- | --- | --- |
| Religious leaders are trusted because they are ingrained in communities and work in communities, unlike politicians | Religious leaders are set apart from politicians because they come from within communities and serve the people | **Establishing religious leaders as non-political actors** |
| People don't listen to political people like they listen to religious people because they understand that when religious people speak it is going to be a message that benefits them. |  |  |
| Messages from religious leaders are taken more seriously because people have more confidence in religious leaders than politicians |  |  |
| Religious leaders have more public confidence than politicians because they can deal with ideologies | Religious leaders are not incentivised by money or other benefits the way politicians are |  |
| People don't trust politicians or medical people so religious leaders are seen as neutral people |  |  |
| People don't have confidence in politicians because they are paid for what they do, unlike religious leaders |  |  |
| Advocating to the government to add a dignified aspect to Ebola burials | Advocating to authorities regarding burial practices | **Establishing confidence and providing community support in the context of an Ebola outbreak** |
| Raising public requests to allow family members to attend Ebola victims' funerals to authorities |  |  |
| Raising public requests to have more women on burial teams to authorities |  |  |
| Providing emotional and spiritual support to people in quarantine | Religious leaders provided support for those in quarantine |  |
| Giving medical advice to people in quarantine |  |  |
| Addressing public fears and concerns regarding burial teams | Religious leaders providing community support for other Ebola related issues |  |
| Support figure for people in communities whose family died from Ebola |  |  |
| Providing support to those who are sick in hospitals and at home |  |  |
| Interreligious collaboration provides a believability and establishes public confidence in religious leaders messages | Interreligious collaboration helped to build confidence in the religious leaders as messengers during the outbreak | **Collaboration built public confidence in religious leaders** |
| Religious leaders speaking with one voice - prevents conflicting messages and increases trustworthiness of the messages |  |  |

|  |  |
| --- | --- |
| Some religious leaders' messages caused confusion early in the outbreak |  |
| Collaboration with technical people regarding specific issues (medical people accompanying to answer medical questions) helps build public confidence in the religious leaders as risk communicators | Collaboration with technical people and local leaders built public confidence in religious leaders messages |
| Collaboration with local leaders and traditional healers to build support in Ebola messages |  |

### Establishing religious leaders as non-political actors

Early in the Ebola outbreak political conspiracies and distrust in political figures were commonplace in Sierra Leone. Rumours circulated among local populations that the government was intentionally spreading the virus along with a general perception of corruption and that the virus was a government ruse to generate international aid (Anderson 2015, Wilkinson 2015). Against this backdrop religious leaders said that they established public confidence as Ebola risk communicators by separating themselves from politicians and political organisations.

> *“…*[the public] *don’t know whether to believe the health workers or whether to believe the politicians, so they were confused. So this made us* [Religious leaders] *to be relevant in the message dissemination because they are seeing us as neutral people.”* (Interview 1 - Muslim)
>
> *“…you will discover that the religious leader will get more results than that politician because they* [politicians] *are coming from the angle of distrust.”* (Interview 4 - Christian)

The religious leaders describe how their separation from political affiliations provided them with a unique platform to approach their risk communication efforts.

> *“Some of them* [the public] *were saying it is good because they lack trust in the politician, but since you people* [religious leaders] *are here, we believe you. We will go by what you say.”* (Interview 4 - Christian)

People within communities recognised the religious leaders when they communicated Ebola messages because, unlike the politicians, they were frequently seen and embedded within communities.

> *“…so that’s the big tool that we have that the politicians don’t have, even those in power. Because maybe he* [Politician] *is in this house* [house belonging to a member of the public]. *The people that are there, he doesn’t know them. A religious person doesn’t do that. I go there first thing in the morning saying “Good morning how is the old man, the old lady, are they okay?” Maybe there is a problem that I need to sort out, to come to their aid.”* (Interview 2 - Muslim)

One religious leader speculated that the reason political figures are not trusted like the religious leaders is because of the financial gain involved in their roles. The religious leaders define their work as vocational and their lack of apparent wealth as a reason to garner public confidence.

> *“When the minister* [political leader] *talks, he is lying. But when I appear on TV…they* [the public] *know when I say something it is going to work, it is going to benefit them. Because they* [the public] *don’t pay me for this work…I don’t have a new bicycle, but I’m dedicated to doing this job”* (Interview 2 - Muslim)

### Establishing confidence and providing community support in the context of an Ebola outbreak

Religious leaders have long provided emotional and spiritual support for many Sierra Leoneans, and they adapted this role to suit the unique needs of their communities during the Ebola outbreak (Featherstone 2015, Hewlett). As a result of the direct and indirect effects of Ebola many Sierra Leoneans found themselves in challenging situations. The religious leaders described themselves as figures of hope for their communities in many of these circumstances, providing a listening ear and spiritual meaning to the situations the public faced.

> *“…we give them the words of hope, in the messages, give them courage that they will not die, they will come out. Let them just listen to what the doctor say. Follow the instructions…. Sometimes they call me 2, 3 am in the morning; “reverend I am giving up, the pain I am feeling so much pain, even to hold the cup for me to drink I cannot, the pain”. I said “This is not for you. You will surely come over this”.”* (Interview 8 - Christian)

This *hope* the religious leaders describe is contextualised in terms of the individual’s spirituality and the essence of placing faith in a higher power beyond the outbreak. The religious leaders describe how they would stand outside the homes of those who were quarantined and talk to them, providing a sense of hope that they will recover from Ebola.

The suffering community members were facing during the outbreak was contextualised by the religious leaders who compared Ebola to the infectious diseases discussed in the Bible (leprosy) and the Qur’an (the*Tha’oon Amwas*). This facilitated public understanding of the Ebola measures religious leaders were communicating while also providing a means to spiritually interpret the outbreak.

> *“There’s nothing in Islam called Ebola. But there are similar things, like the tha’oon* [infectious disease discussed in the Qur’an]*”* (Interview 2 - Muslim)
>
> *“In the bible lepers were set apart…because they thought it was a contagious disease”*
>
> (Interview 9 - Christian)

The religious leaders further report building confidence in their messages by advocating for the needs of their communities. Many Sierra Leoneans were frustrated by the specific restrictions placed upon them during the outbreak, for instance, restrictions on traditional burial practices which were imposed by the government to reduce the spread of Ebola (Jalloh 2020). These restrictions did not consider the spiritual and cultural needs of the public. Religious leaders worked with (medical) authorities to design safe and dignified burials, including incorporating more women into burial teams in response to concerns that deceased women were being buried by men.

> *“The burial team was full of male. So at that time, the women, they rose up and they said uh no! We need women in the burial teams. We went to the government and we said this is the concern of our people….The government agreed.”* (Interview 4 - Christian)

The religious leaders described this advocacy as a tool to provide their communities with what they needed at a time of crisis and in doing so built trust in their role as communicators.

> *“* [the public] *want to appeal to you, to appeal to the government in the area of burial…even though they are talking about safe burial, we want to add dignity into it. It is our culture, we need to give our funeral rights to our loved ones when they die.”* (Interview 8 - Christian)

### Collaboration built public confidence in religious leaders

During the Ebola outbreak Christian and Islamic leaders worked together to develop and communicate Ebola messages across Sierra Leone in a coordinated manner. The religious leaders described how this interreligious collaboration was beneficial for the risk communication campaigns as it emphasised, for the public, the importance of putting aside religious differences to focus on the outbreak.

> *“At first, they* [the public] *were thinking that because of our different beliefs it would be difficult for us* [Muslims & Christians] *to come together, you know, but we were able to make the people know that, no, in fact, it is the source of our strength to come together… Some of us were able to say look, when Ebola is coming it is not looking for a Christian, it’s not looking for a Muslim, it’s not looking for your tribe, it’s not looking for your colour, it is just looking for an individual that it can suppress, kill, destroy.”* (Interview 4 - Christian)

There was a positive public reaction to seeing Islamic and Christian leaders entering towns and villages together according to the religious leaders. They describe the positive effect of seeing religious leaders standing side-by-side in Mosques and Churches communicating in unison.

> *“We do everything in common. Sometimes we even go into churches, when it is required to go to the church, we go to the church when it is required to come to the mosque, we do that also. Sometimes you can see some Christians in the mosque, sometimes you can see some Muslim in the church. Just for the people to get the message.”* (Interview 1 - Christian)

This interreligious collaboration was described from a more pragmatic perspective by a number of the religious leaders. They acknowledged the confusion that different, often conflicting messages from multiple sources caused during the outbreak. They considered religious leaders of different doctrines speaking with one unified voice as much clearer and more inherently understandable to their audiences.

> *“…you know initially, at first, the misinformation was so much…the information was coming in every direction….everyone was confused.”* (Interview 6 - Christian)
>
> *“…we said that we are going to speak with one voice. We don’t want to speak with conflicting messages. So that at least when you go to the south, I go to the north and he goes to the east, we make sure that the message that you are giving the people is the same message. Don’t say any different thing. So, we all speak with one voice. Yes, no conflicting message.”* (Interview 7 - Christian)

There was acknowledgement from the religious leaders that they contributed to the public’s confusion early in the outbreak. The subsequent unified approach to risk communication was described as important to prevent false information being spread by religious leaders.

> *“…messages came that people should wash with water and salt, then the Ebola will not affect you, oh yeah…a prophet from Nigeria, you know, they gave a portion* [of the bible]*… it went like a wildfire. Even Muslims were washing, they were bathing with that stuff”* (Interview 4 - Christian)

Collaboration was also described by the religious leaders in terms of their interactions with other sectors of the Ebola response and the beneficial impact this had on their work overall. When communicating their messages, the religious leaders often entered communities as part of a larger team which included medical experts and other non-religious people. The religious leaders’ presence beside these medical teams made their messaging more acceptable to the public.

> *“When it comes to the technical parts of the disease, we call on the technical people*. *Because when we are going* [into communities] *sometimes we go with some health officers, we involve the DHMT* [District Health Medical Team]*… We will call them to come on board and they explain to the people. But because we are around, they* [public] *will accept them* [medical team messages]*.”* (Interview 4 - Christian)

The religious leaders would also engage village chiefs, community leaders and traditional healers upon entering towns and villages as a means to support their communication campaigns. These local leaders would help to disseminate messages through the communities using megaphones and going house to house to spread the messages. The religious leaders also described their approach to collaboration in pragmatic terms. Although they may not have agreed with external parties such as the traditional healers, they were willing to put this aside for the greater good. They described these actions in terms of how best to communicate their messages.

> *“We also saw it necessary to work with the traditional healers because the practice of the traditional healers very contradicts the belief of the Christian and the Muslim. But because it was a national emergency the thing* [Ebola] *was killing our people, we had to say no, let’s ignore some of this…fundamental right…and then address the issue that is killing people.”* (Interview - Christian)

## Discussion

Three primary strategies were described in the data which religious leaders used to build confidence in their role communicating risk and the messages they communicated; establishing themselves as non-political actors in the outbreak, providing community support and using collaboration to build confidence in their communicated messages. They established new strategies as well as building upon pre-existing religious roles in Sierra Leonean society to achieve this. Religious leaders pragmatically utilised scripture to communicate risk and also to establish confidence in their messages.

Trust in institutions and governments is considered a key component of an effective outbreak response (36). Our findings suggest that religious leaders built confidence in their messages by distancing themselves from politicians, who were not widely trusted (37, 38). With effect it seems, as public health messages promoted by religious leaders have also been positively associated with willingness to engage in safe burial behaviours during the Ebola outbreak in Sierra Leone (21). Instead of associating with political leaders, the religious leaders sought different trust alliances – for instance through inter-religious collaborations.

While Sierra Leone is unique with its high levels of interreligious tolerance, the power of communicating with a unified message holds lessons for other emergency settings. Even though the leaders in this study did not discuss the ideological conflicts which can exist between the two religions (39), our findings highlight the importance of finding a middle ground to help bolster a collaborating position. This is also shown through the religious leaders’ collaboration with traditional healers, for which doctrinal differences were put aside to fight their common enemy. The religious leaders seem to appreciate the positive impact that seeing others cooperate can have on an individual’s willingness to cooperate (36, 40).

A number of the religious leaders in our study referred to themselves as *non-political* and *neutral* when describing their role in Sierra Leonean society. While they pragmatically separated themselves from political affiliations to assist in their risk communication strategies, evidence suggests that religion and politics are closely intertwined and difficult to separate from one another in Sierra Leone (23, 33). The religious leaders in our study do not reflect upon this when referring to their role as *non-political* and *neutral.* Similarly, they do not reflect upon some of their religious doctrines which are in conflict with basic public health messages such as contraception and birth control which have been linked both positively and negatively to religious involvement (9–11, 13). While religious leaders were associated with uptake of positive health behaviours during the Ebola outbreak in Sierra Leone the influence these roles garner may well carry over to other less favourable impacts on public health and political influence which needs to be considered when looking towards ongoing and future outbreaks.

Religious leaders were embedded in communities and could appreciate the specific needs of the individual community members while also distributing messaging through wider communication campaigns. The socio-ecological framework highlights the importance of intrapersonal and interpersonal relationships when trying to affect behaviour change (17, 41). The top-down approach to communication seen early in the outbreak was not as effective in part because it did not account for the socio-cultural realities of many Sierra Leoneans. For example, burial teams were not always dignified making accepting them challenging for many, and avoidance of caregiving for sick family members was not always an option (1, 42).

The influence of socio-ecological factors on the uptake of protective health-behaviours was similarly demonstrated during the COVID-19 pandemic. Adherence to mask wearing guidance and compliance with self-isolation were positively associated with household income, age, personal attitudes, educational attainment and gender (43, 44). The COVID-19 pandemic has demonstrated the important role of trusted actors in the public health response, including religious leaders (45). The importance of building public confidence and trust in the messenger is key to effective risk communication as has been established previously using the socio-ecological model (36, 46, 47).

Religious leaders describe their scriptural texts as an integral part of risk communication campaigns. From contextualising the outbreak for those who are sick and fearful, to developing the risk communication messages around scripture, the religious texts were ingrained across their actions. Producing messages in support of public health messages adapted to suit the needs of a population can help acceptability and efficacy of public health programmes (48, 49). A willingness to engage with the medical professionals and build Ebola specific messages from scripture may be a practical and beneficial application of the findings of this study.

## Strengths and limitations

The religious leaders in this study are a diverse cohort of participants with male and female, Christian and Islamic leaders represented. The study sample is limited to the perspective of religious leaders, most of whom held powerful positions in religious and faith-based organisations in Sierra Leone. As such, their opinions may not be entirely representative of all religious leaders in Freetown and indeed those in more rural parts of the country. Nonetheless, the perspective of these leaders may give a more overarching viewpoint of religious leader involvement in the outbreak.

While the sample was limited to 10 religious leaders, the power could be considered robust since the aim of the study was well-focused, the sample of participants was unique and the participation rate was 100% (50). The study was also informed by theory and the participants provided in depth and reflective responses.

A few potential sources of bias should be mentioned. Interviews were carried out by PL who is a caucasian male, which may have introduced a response bias. Arguably the religious leaders may have provided more candid responses to a Sierra Leonean interviewer. On the other hand, they may have provided the most honest answers to someone they perceive as outside the local healthcare system particularly when criticising the outbreak response efforts. Harvey’s “elite interviews” also helped to inform PL on how to approach the interviews in terms of building trust and engaging these religious leaders who are considered high standing community figures (51). Participants were all interviewed in Freetown. The experience of the outbreak may have been significantly different for those living in remote and rural areas and may differ between regions, therefore findings are limited to Freetown. Eight out of 10 of the interviews were conducted in FOCUS1000 headquarters, potentially leading to a response bias. This effect was somewhat mitigated by reiterating that PL was not affiliated with FOCS1000 and all interviewees’ responses would remain entirely confidential. Finally, an unavoidable source of bias in this study was that the qualitative interviews were conducted more than three years after the outbreak, which might have led to omission of details and possible recency bias in terms of some of the later successes seen in the outbreak.

## Conclusion

Religious leaders saw themselves as key actors in risk communication campaigns during the Ebola outbreak in Sierra Leone. They responded to the needs of their communities and the rapidly changing context of the outbreak to communicate risk. In this study religious leaders highlighted the key role that inter-religious collaboration and collaboration with medical experts had in their role communicating risk and applied their scriptural texts to suit the needs of the outbreak response. They were pragmatic in their approach to risk communication and engaged with multiple different facets of the outbreak response to strengthen their position. Religious leaders are embedded across Sierra Leonean society within small communities through to nationwide organisations. It is these broad ranging networks that allow religious leaders to effectively approach risk communication strategies at multiple levels while accounting for the socio-ecological realities of the individual.

## Declarations

### Ethics approval and consent to participate

The Sierra Leone Research and Scientific Review Committee granted ethical approval for this study in March 2019. All interviewees provided informed consent to participate by signing an informed consent form.

## Consent for publication

No consent was required for publication, as no identifiable information is contained within.

## Availability of data and materials

The datasets used and/or analysed during the current study are available from the corresponding author on reasonable request.

## Authors’ contributions

PL, MW, MFJ, and HN led the overall study design with substantial contributions made by all co-authors for one or more of the study components. PL organised and coordinated primary data collection with assistance from FOCUS100. PL led secondary data analysis with support from MW, HMA & HN. PL, MW, HMA and HN provided critical input to the data interpretation. PL led the writing of the manuscript with substantial contributions made by all co-authors. All the authors reviewed and approved the final version of the manuscript.

## Data Availability

The underlying data will be made available from the authors on reasonable request.

## Acknowledgements

The authors would like to thank Mr Mohammad B Jalloh, Mr Paul Sengeh and everyone at FOCUS1000 in Sierra Leone for their collaboration on this research project and their tireless work during the pandemic. The authors would also like to thank the religious leaders for their participation in the study.

